# Prediction of Pivot Shift Grade Using In-Vivo Ultrasound Bone Tracking During Sit-Stand-Sit: A Machine Learning Feasibility Study

**DOI:** 10.64898/2026.05.10.26352712

**Authors:** Jiya Dutta, Isaac Tay, Lai Kah Weng, Jeremy Lim Tze En, Chia Zi Yang

**Author notes:** **Corresponding Author:** *Jiya Dutta.

## Abstract

**Background:** The pivot shift (PS) test is the most specific clinical examination for anterolateral rotational instability in ACL-deficient knees, yet grading remains subjective, as evidenced by poor inter-observer reliability, particularly for Grade 2. Since low-grade (Grade 1) versus high-grade (Grades 2/3) PS is the threshold for recommending lateral extra-articular augmentation, performing the test in awake clinic patients limits grading reproducibility and introduces variability in surgical decision-making. Existing methods to quantify the pivot shift usually require examiner-performed testing under general anaesthesia. No prior approach has ascertained PS grading from a separate patient-performed functional movement.

**Purpose:** To evaluate the feasibility of a machine learning (ML) classifier, trained on kinematic ultrasound bone-tracking signals acquired during a patient’s sit-stand-sit (SSS) knee movement, to predict their PS grade, and to clinically validate its ability to differentiate low versus high-grade PS.

**Methods:** Ultrasound bone-tracking kinematic data were collected during SSS manoeuvres in 23 ACL-injured patients using the GATOR device, and ground truth PS grades (0–3) were assigned under general anaesthesia by fellowship-trained orthopaedic sports surgeons. From the data collected, Leave-one-out cross-validation (LOOCV) was used to train the ML classifier. Clinical SSS data from 6 ACL-deficient patients was used for independent held-out validation of their low-grade (Grade 1) versus high-grade (Grade 2/3) PS.

Multiple deep learning architectures (XceptionTime, InceptionTime, FCN, ResNet, ResCNN) and training strategies (including mixup augmentation and supervised contrastive learning) were tested. Performance was measured by one-versus-rest (OVR) AUC under LOOCV and by AUC (low vs high grade PS) from the held-out patient sessions.

**Results:** The ML classifier achieved a maximum OVR AUC of 0.928 ± 0.084 under LOOCV. Classifier performance increased with pivot-shift severity: Grade 3 was identified most reliably (AUC ~0.81; sensitivity 0.70–0.80), whereas Grade 2 remained the most challenging boundary (sensitivity 0.20–0.75 across configurations). For the clinically relevant binary classification of low-versus high-grade pivot shift, the classifier generalised well to a completely unseen patient cohort (AUC 0.889; accuracy 0.860; sensitivity 0.850; minimum-class sensitivity 0.767).

**Conclusion:** The study demonstrates that kinematic ultrasound bone-tracking during sit-stand-sit contains transferable information about rotational instability severity in ACL-deficient patients, and represents the first reported approach to predict pivot shift grade from a patient-performed functional movement. The strong cross-validation performance confirms that the signals contain meaningful PS grade-discriminative information, but larger datasets targeting 50–100 sessions per grade will be required to achieve patient-level generalisation and advance this novel rotational instability assessment tool toward full clinical adoption.

**Level of Evidence:** Level IV, diagnostic feasibility study.

## Introduction

The pivot shift (PS) test is widely regarded as the most specific clinical examination for detecting and grading anterolateral rotational instability of the knee due to an anterior cruciate ligament (ACL) tear or deficiency [1]. Higher pre-operative PS grades have been independently associated with increased rates of graft revision [2], while post-op residual pivot shift has been linked to early knee osteoarthritis [3].

The PS grade serves as an important clinical data point for surgical decision-making, with patients showing Grade 2 or 3 instability more likely to benefit from augmentation of standard ACL reconstruction with a lateral extra-articular procedure (LEAP), as demonstrated by the STABILITY trial [4]. However, the decision for lateral augmentation is weighed together with other patient-specific factors, including age, activity level, sport demands, concomitant injuries, and surgeon preference.

Many surgeons continue to perform the PS test routinely in the awake outpatient setting despite its poor inter-observer reliability and subjectivity [5], as no better method currently exists to assess dynamic rotational instability. During examination under general anaesthesia (EUA), the PS test benefits from full muscle relaxation, providing more consistent evaluation of dynamic rotational laxity. Traditional quantitative research efforts—including computer navigation systems, inertial sensors, smartphone accelerometry, and optical motion capture—rely on kinematic signals recorded during an examiner-performed manoeuvre and most achieve reliable accuracy only under EUA [6–8].

Because most of these approaches require EUA, quantitative PS devices are largely confined to the perioperative period. For outpatient assessment of dynamic knee rotation in awake patients, measurement devices are compromised by skin artifact errors, which B-mode ultrasound bone surface tracking have been shown to be able to overcome [9,10].

The GATOR (PreciX Pte Ltd, Singapore) is a non-invasive real-time ultrasound bone-tracking device that measures dynamic knee rotational laxity during a patient-performed SSS task. It is combined with inertial sensors [11] to overcome skin motion artifacts and accurately quantify femur-on-tibia knee rotation [12].

As machine learning (ML) techniques have recently been used to model nonlinear interactions within multidimensional clinical datasets including ultrasound [13], we hypothesised that the rotational kinematic signature captured by ultrasound bone-tracking during SSS movement would contain sufficient discriminative information to allow an ML classifier to predict the PS grade.

This study evaluates the feasibility of an ML classifier trained on kinematic ultrasound bone-tracking signals acquired during patient-performed sit-stand-sit to predict pivot-shift grade (0–3) and to determine whether the clinically relevant distinction between low-grade (Grade 1) and high-grade (Grades 2/3) instability generalises to a completely unseen patient cohort.

## Methods

### Patients and Data Collection

This was a single-center prospective cohort study conducted between January 2019 and December 2024 at Singapore General Hospital (SGH). Ethical approval was obtained from the SingHealth Centralized Institutional Review Board (CIRB 2019/2766), and the study was conducted in accordance with PDPA regulations. All participants provided written informed consent. At the interim analysis, 29 patients had been recruited.

Eligible patients were adults scheduled for primary ACL reconstruction with preoperative MRI confirmation of a complete ACL tear. Exclusion criteria included multiligamentous knee injuries, revision ACL procedures, previous contralateral ACL reconstruction, and any prior knee surgery.

For ground truth labelling of training data set, kinematic ultrasound bone-tracking data were successfully acquired from both knees of 23 distinct patients during SSS maneuvers using the GATOR device, as bilateral data collection was integral to the analysis design. Fellowship-trained orthopaedic sports surgeons with substantial expertise in knee instability assessment assigned the Pivot shift grading (Grade 0: absent; Grade 1: glide; Grade 2: clunk; Grade 3: gross subluxation) under anesthesia [14].

From the prospectively recruited cohort at SGH, six ACL-deficient patients were allocated to the held-out validation test set.

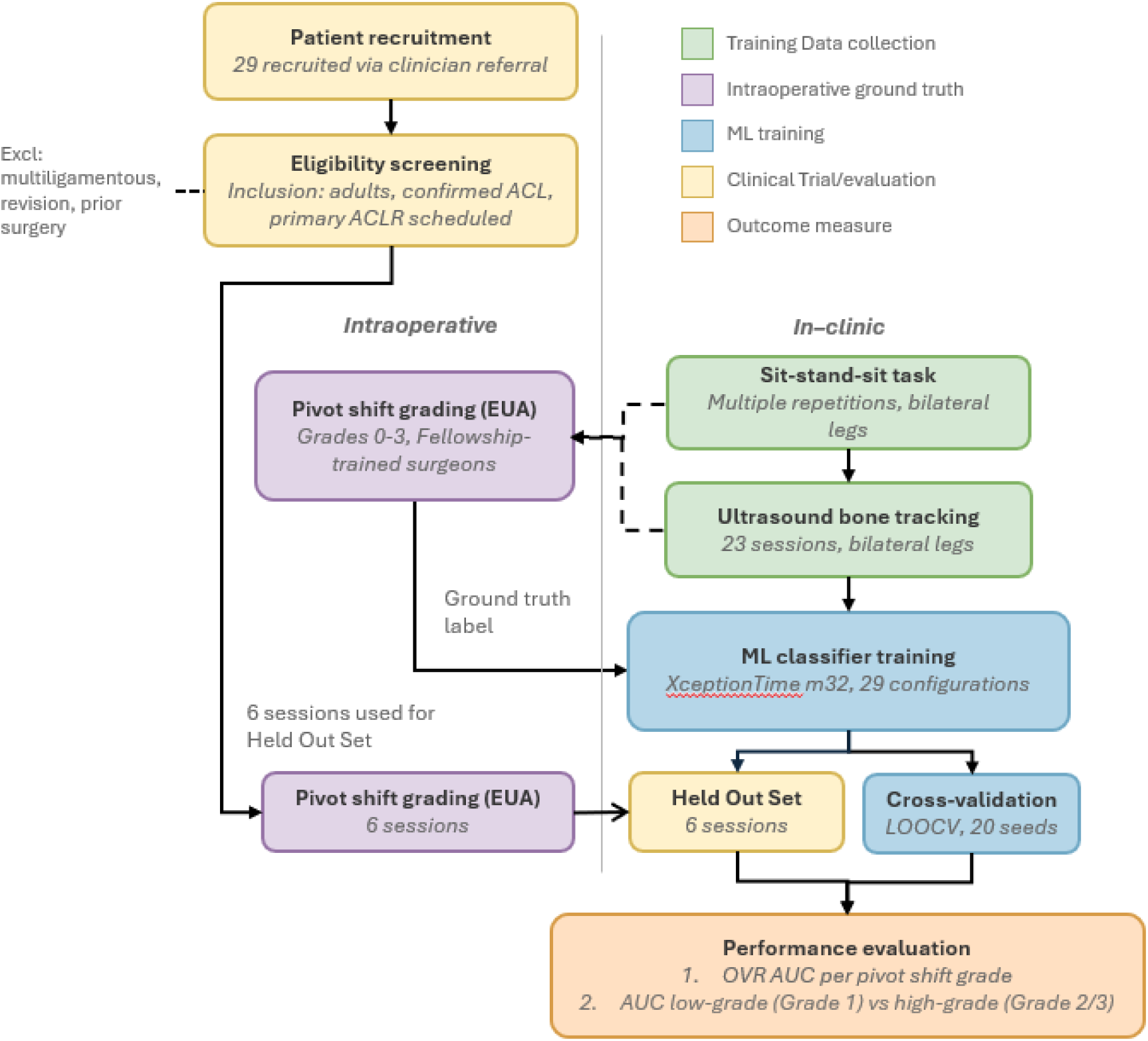

The SSS task was selected for its biomechanical relevance: the extension phase loads the knee through the screw-home rotation arc, engaging the screw-home mechanism that is disrupted in ACL-deficient knees [16].

### Statistical Analysis

Performance was measured using the area under the receiver operating characteristic curve (AUC), a standard diagnostic accuracy metric where 1.0 represents perfect discrimination and 0.5 represents chance. AUC was evaluated separately for each PS grade against all others (one-versus-rest: OVR AUC). All metrics are reported as mean ± standard deviation across seeds. Leave-one-out cross-validation (LOOCV) was used on the 23-patient dataset.

To align the analysis with a surgically actionable decision - i.e. LEAP augmentation, we additionally evaluated a binary low-versus-high grade formulation, in which Grade 1 (glide) was compared against the merged Grade 2 (clunk) & 3 (gross subluxation) group [17]. This threshold reflects a clinically meaningful decision point in ACL care: high-grade pivot shift is a recognised indication for considering adjunct lateral extra-articular augmentation. Generalisation to completely held-out patient sessions (the 6 withheld patients) was assessed as the primary clinical validity criterion, with the binary low-versus-high grade held-out AUC serving as the clinically-relevant endpoint.

## Results

### Grade-Dependent Kinematic Signatures

The kinematic ultrasound bone-tracking signal during sit-stand-sit (SSS) contains grade-discriminative information that reflects PS severity. Performance followed a clear monotonic relationship with laxity severity. Grade 3 (gross subluxation) was the most reliably detected (AUC ~0.81), while Grade 0 was the most difficult to isolate. Grades 1 and 2 showed intermediate performance, with Grade 2 sensitivity ranging from 0.20 to 0.75 across methods.

**Figure 1.**
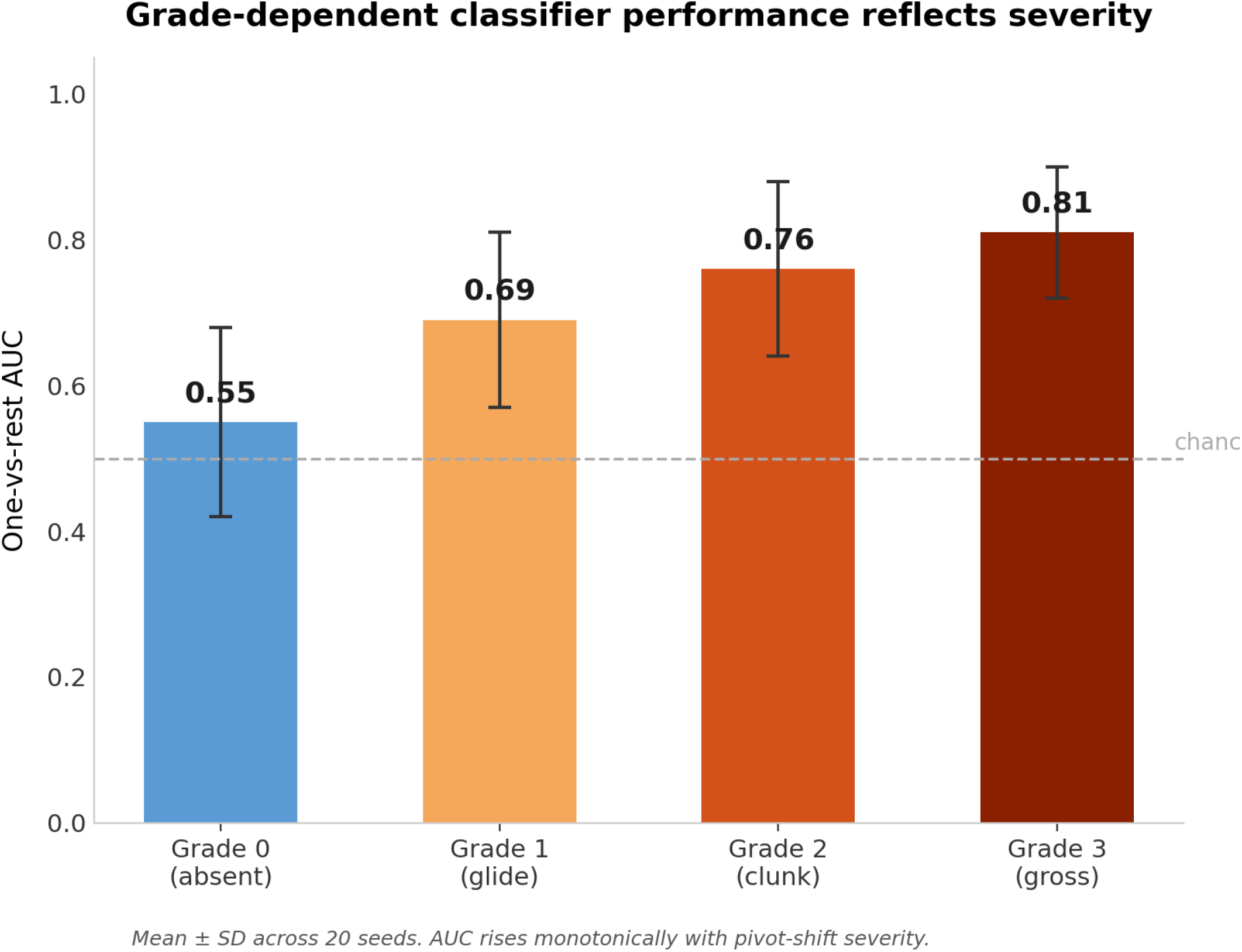
OVR AUC across different grades reflecting monotonic rise with severity.

**Table 1.**
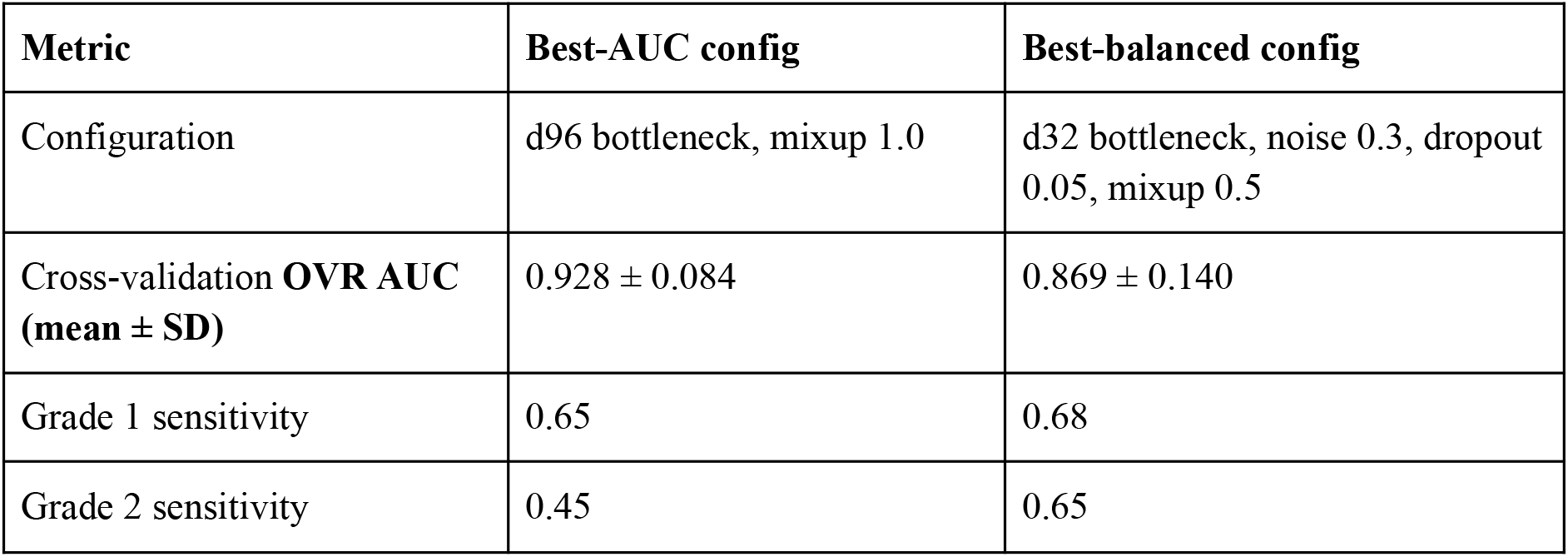
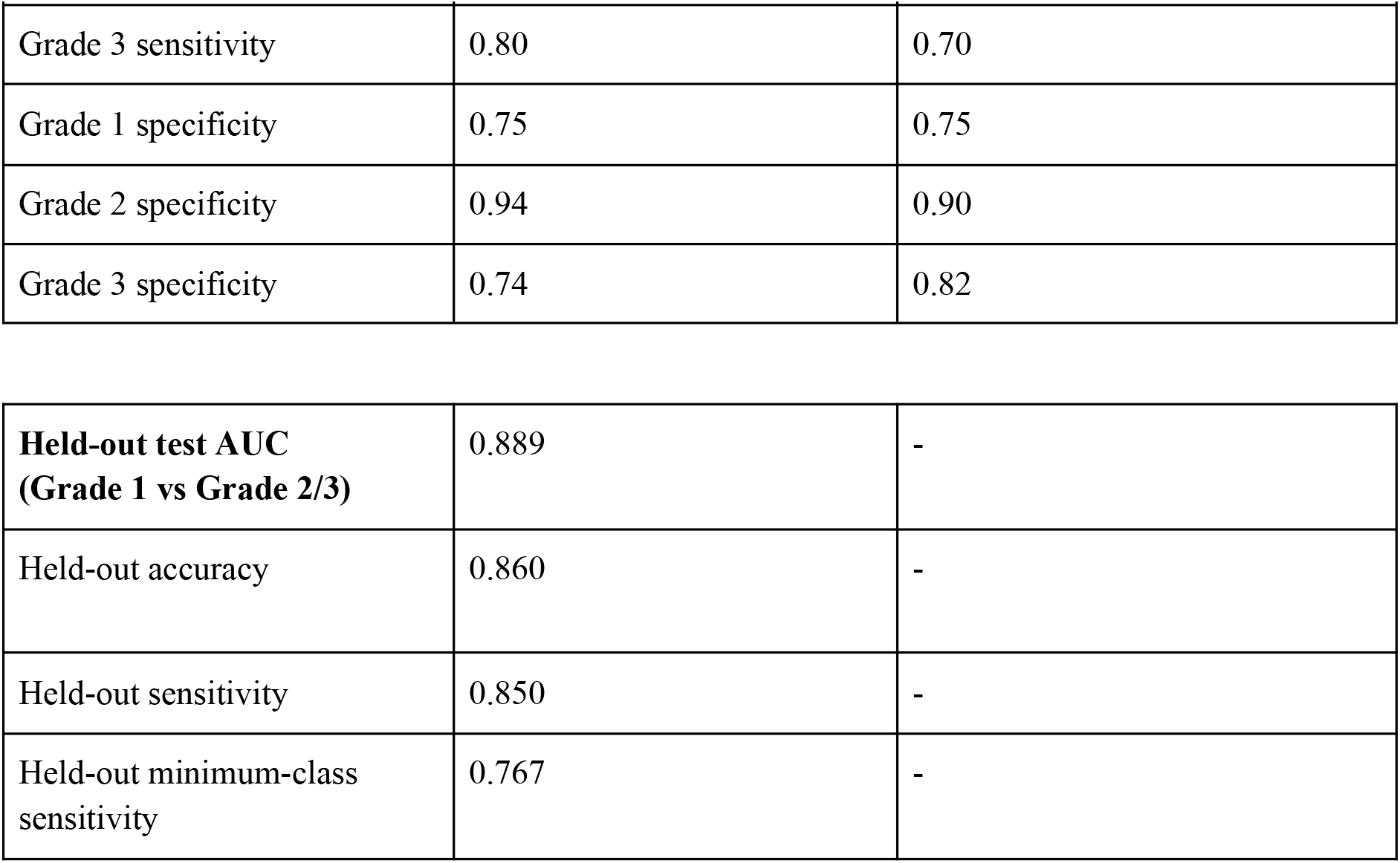
Cross-validation and held-out performance of the two reference configurations.

| Metric | Best-AUC config | Best-balanced config |
| --- | --- | --- |
| Configuration | d96 bottleneck, mixup 1.0 | d32 bottleneck, noise 0.3, dropout 0.05, mixup 0.5 |
| Cross-validation **OVR AUC** (mean ± SD) | 0.928 ± 0.084 | 0.869 ± 0.140 |
| Grade 1 sensitivity | 0.65 | 0.68 |
| Grade 2 sensitivity | 0.45 | 0.65 |
| Grade 3 sensitivity | 0.80 | 0.70 |
| Grade 1 specificity | 0.75 | 0.75 |
| Grade 2 specificity | 0.94 | 0.90 |
| Grade 3 specificity | 0.74 | 0.82 |
| **Held-out test AUC**<br>**(Grade 1 vs Grade 2/3)** | 0.889 | - |
| Held-out accuracy | 0.860 | - |
| Held-out sensitivity | 0.850 | - |
| Held-out minimum-class<br>sensitivity | 0.767 | - |

### Overall Classifier Performance and Generalisation

On the clinically actionable threshold - a binary low-grade (Grade 1) versus high-grade (Grade 2/3) classification - the classifier generalised well, with an AUC of 0.889, an accuracy of 0.860, sensitivity of 0.850, and a minimum-class sensitivity of 0.767. This binary distinction helps a surgeon with offering a patient lateral extra-articular tenodesis to the ACL reconstruction. On the 6 held-out patients, the model correctly classified the low vs high grade PS 86% of the time (accuracy 0.860).

When it predicted high-grade instability, it was right 85% of the time (sensitivity 0.850). Importantly, the classifier demonstrated balanced performance without systematic bias toward either category. It performed comparably across both low- and high-grade pivot shift groups, achieving a minimum sensitivity of 0.767. This confirms that the model did not simply classify all knees as high-grade to artificially improve performance metrics. A surgeon seeing a new ACL patient in the clinic for the first time could expect the system to correctly identify whether that patient has high-grade rotational instability approximately 9 times out of 10. That is the threshold distinction that would inform whether a lateral extra-articular procedure is indicated [4].

## Discussion

The clinical motivation for this work stems from a persistent and well-recognised gap in the ACL care pathway. While anterior-posterior laxity can be reliably quantified with instruments such as the KT-1000 arthrometer and structural integrity can be assessed by MRI, dynamic rotational instability continues to be evaluated almost exclusively by subjective clinical testing.

This feasibility study demonstrates that ultrasound bone tracking during a patient-performed sit-stand-sit (SSS) task captures quantitative kinematic information that correlates with pivot shift (PS) grade in ACL-deficient knees. Importantly, the bone movement signatures showed a clear monotonic relationship with clinical laxity severity: discrimination performance improved progressively from Grade 0 to Grade 3. Grade 3 (gross subluxation) produced the most distinct and reliably identifiable kinematic pattern (OVR AUC ~0.81), whereas Grade 2 remained the most challenging boundary. This pattern closely mirrors the well-documented poor inter-observer reliability of manual PS grading in the clinical literature, particularly at the Grade II threshold [5]. These findings provide the first objective evidence that a simple, examiner-independent functional movement can encode meaningful information about rotational instability severity.

The pivot shift grade is a critical decision-making variable — Grade 2 or 3 instability is the key indication for lateral extra-articular augmentation in many contemporary protocols [4] — yet its assessment in awake patients remains highly variable. The present approach is explicitly designed to complement, rather than replace, existing clinical evaluation. This interpretation is consistent with recent ML-based work on postoperative rotational instability prediction [15]. By providing an objective, quantitative kinematic measurement during a standardised weight-bearing task that patients can perform independently, ultrasound bone tracking has the potential to bring reproducible rotational laxity assessment into routine outpatient, rehabilitation, and return-to-sport settings.

The choice of the sit-stand-sit movement is biomechanically grounded. Unlike examiner-driven pivot shift manoeuvres, the SSS task is powered entirely by the patient’s own musculature and body weight. During the terminal extension phase, the knee naturally engages the screw-home mechanism — the obligatory external tibial rotation that occurs with full extension. In ACL-deficient knees this rotational coupling is disrupted, resulting in anterior and internal tibial subluxation followed by abrupt reduction [16]. The GATOR system’s ability to track underlying bone surfaces in real time allows these subtle kinematic events to be captured with high precision, free from the skin-motion artefact that limits conventional marker-based or inertial systems in awake patients.

To our knowledge, this is the first study to attempt prediction of PS grade from a completely separate patient-performed functional movement without replicating the examiner-driven pivot shift test itself [6–8,16]. Previous machine-learning efforts have focused almost exclusively on quantifying the pivot shift manoeuvre performed under anaesthesia [6–8]. The present work therefore represents a conceptual advance toward truly examiner-independent, point-of-care rotational instability assessment.

The grade-dependent pattern of classifier performance warrants a mechanistic explanation that goes beyond sample size. Two distinct mechanisms explain this pattern: First, Grade 3 benefits from a signal magnitude advantage - gross subluxation produces a large-amplitude, abrupt bone displacement event during the screw-home arc that is sufficiently large to be differentiated from Grades 1 and 2 even with very few training examples. The classifier does not need a large dataset to learn a feature that is biomechanically obvious and discernible. Second, the Grade 1/Grade 2 boundary represents a signal separability problem that is largely independent of dataset size. The distinction between these grades is subtle enough that neither the current classifier nor clinical examiners can resolve it reliably — reflected in the κ = 0.06 inter-observer agreement for Grade 2 [5]. Increasing the dataset will reduce estimation variance at this boundary but will not resolve the underlying ambiguity without task modifications or additional signal modalities that amplify the difference between Grades 1 and 2. Future work should therefore pursue both dataset expansion and investigation of task or sensor modifications that improve the Grade 1/Grade 2 signal separability that constrains classification performance independently of sample size.

For performance analysis of the clinical unseen cohort, the classification task was reframed to a binary distinction - by collapsing the four-grade PS scale into low-grade (Grade 1: glide) versus high-grade (Grades 2/3: clunk or gross subluxation), which is the established threshold for recommending LEAP adjunct to ACL reconstruction. By collapsing Grades 2 and 3 into a single high-grade category, the ML classifier is no longer required to resolve the subtle and challenging distinction at the Grade 1/Grade 2 boundary. Instead, it separates the diffuse low-grade glide pattern (Grade 1) from a combined class defined by clinically significant subluxation (Grades 2/3). This reformulation produces a clearer kinematic separation, resulting in strong generalisation to the held-out test set (AUC 0.889, accuracy 0.860, sensitivity 0.850, minimum-class sensitivity 0.767). These findings demonstrate that kinematic ultrasound bone-tracking signals acquired during sit-stand-sit contain transferable, patient-independent information about the surgically actionable degree of rotational instability, even when the dataset remains too small to resolve all four ordinal grades on completely unseen patients.

In summary, this feasibility study provides encouraging proof-of-concept that ultrasound bone tracking during the sit-stand-sit manoeuvre can extract clinically relevant information about rotational knee instability. The kinematic signals showed a positive relationship with pivot-shift severity, performing better as laxity increased, and most-reliably identified Grade 3 instability. The clinically relevant distinction between low- and high-grade pivot shift was able to generalise to unseen patients, even at the current dataset size (AUC: 0.889). These findings establish that the sit-stand-sit bone-tracking signal contains the biomechanical information required for objective rotational laxity assessment, from which larger prospective validation studies can further refine the ML classifier. Future work should focus on expanding the dataset, standardising the sit-stand-sit protocol, incorporating healthy controls, exploring hybrid models that integrate kinematic features with additional clinical and imaging data, and investigating task or sensor modifications that improve Grade 1/Grade 2 separability.

## Limitations

This study has several important limitations that should be considered when interpreting the results. The primary limitation is the small sample size with respect to generalisation. Only 23 patient sessions were available for leave-one-out cross-validation, with just six additional sessions reserved for independent held-out validation. This number is insufficient for robust multiclass deep learning classification across 4 pivot shift grades. It is important to note, however, that sample size is not the sole explanation for all performance limitations observed. The Grade 1/Grade 2 classification challenge reflects a fundamental signal separability problem that is subtle enough to challenge both the classifier and experienced clinical examiners alike — reflected in the κ = 0.06 inter-observer agreement for Grade 2 — and this boundary is unlikely to be fully resolved by dataset expansion alone without complementary changes to the task design or sensor configuration. Importantly, the held-out binary result demonstrates that this ambiguity does not preclude clinically useful generalisation: by aligning the classification target with the surgically actionable low-versus-high grade threshold, the classifier was asked to separate a diffuse glide pattern from a class defined by clinically significant subluxation — a kinematically larger and more consistent separation that transferred to entirely unseen patients.

In addition, the sit-stand-sit (SSS) protocol was not fully standardised across participants with respect to chair height, movement speed, foot positioning, or arm usage. These variations likely introduced additional kinematic noise unrelated to rotational instability, which may have reduced the signal-to-noise ratio and contributed to the observed generalisation challenges.

Another key limitation relates to the ground truth labels. Pivot shift grades were determined under general anaesthesia by experienced fellowship-trained surgeons, which remains the current clinical gold standard. However, the degree to which bone kinematics captured during a voluntary, weight-bearing sit-stand-sit movement in awake patients correlate with the passive examiner-driven pivot shift manoeuvre performed under anaesthesia remains unknown. This potential mismatch between testing conditions represents an important area for future validation.

A further limitation concerns the assumption that pivot shift grade reflects a clean, isolated measure of rotational laxity in all patients. In practice, ACL-deficient knees frequently present with concomitant pathology — including meniscal tears, chondral lesions, posterolateral corner insufficiency, and anterolateral ligament injury — that may independently contribute to or modify the rotational kinematic signature captured during sit-stand-sit, independently of the assigned PS grade. Similarly, the contralateral limb, while clinically considered the reference healthy knee, cannot be assumed to be biomechanically normal in all cases. Subclinical laxity, previous injury, early degenerative change, or constitutional ligamentous laxity may cause the contralateral knee to exhibit a non-zero PS grade or an elevated rotational range, meaning that side-to-side comparisons used to contextualise the injured limb’s kinematics may not always reference a truly normal baseline. These factors introduce variability in both the labelling and the bilateral reference signal that are difficult to control for at this dataset size, and future studies should systematically document and account for concomitant pathology and contralateral knee status in their analysis design.

Finally, the study cohort consisted exclusively of patients with confirmed ACL injuries undergoing reconstruction. The absence of a healthy control group limits the ability to establish baseline performance for binary (injured vs. uninjured) detection and to fully characterise normal kinematic variability during the SSS task. Inclusion of asymptomatic controls in future studies would strengthen the interpretation of the grade-specific signatures observed here.

Despite these limitations, the promising cross-validation results, together with held-out generalisation at the surgically actionable low-versus-high grade endpoint, provide proof-of-concept that ultrasound bone tracking during a patient-performed functional task contains clinically relevant information about rotational instability severity.

## Conclusion

This feasibility study demonstrates that kinematic ultrasound bone tracking during SSS captures quantitative information that reflects the severity of rotational knee instability in ACL-deficient patients. The magnitude and distinctiveness of grade-dependent bone movement signatures increases with PS severity, with Grade 3 instability producing the most reliably identifiable signal. The discrimination of low-grade (Grade 1) from high-grade (Grade 2/3) pivot shift - the threshold at which lateral extra-articular augmentation of ACL reconstruction is indicated - did generalise with a held-out AUC of 0.889. Expanded prospective data collection, targeting a minimum of 50--100 sessions per grade, is required to extend full ordinal grading and to confirm the clinical validity of the low-versus-high grade discrimination at scale.

## Data Availability

All data produced in the present study are available upon reasonable request to the authors.

